# Characterizing Parkinson’s Disease Clinical and Biomarker Interactions in REM Sleep Behavior Disorder

**DOI:** 10.1101/2025.05.16.25327469

**Authors:** Vijaya L. Reddy, Samantha Esposito, Erika Renkl, Amine Benyakoub, Kara Mead, Camalene Chrysostoum, Sapna Patel, John P. Seibyl, Yuan Huang, Brian B. Koo, Jesse M. Cedarbaum

**Author notes:** **Corresponding Author Contact Information:** Name: Jesse M. Cedarbaum MD, Address: 300 George St. Suite 353, New Haven CT 06511.

## Abstract

**Background:** REM Sleep Behavior Disorder (RBD), marked by dream enactment due to loss of REM-related muscle atonia, is a prominent prodromal indicator of synucleinopathies, particularly Parkinson’s Disease (PD).

**Objectives:** This study aimed to investigate the interplay among key PD biomarkers— α-synuclein seed amplification assay (SAA), hyposmia, and dopamine transporter (DaT) SPECT imaging — in individuals with RBD. Additionally, we evaluated how phenoconversion events and Movement Disorder Society (MDS)-Prodromal PD probability scores relate to clinical symptoms and biomarker profiles in an incident RBD population.

**Methods:** Participants with polysomnographically-confirmed RBD underwent comprehensive clinical and biomarker assessments. They were grouped along three non-exclusive biomarker-based axes (hyposmic vs. normosmic, SAA positive vs. SAA negative, and DaT positive vs. intermediate vs. negative) and two clinical outcome-based axes (high vs. intermediate/low MDS-Prodromal PD probability; phenoconverters vs. non-phenoconverters). Within each category, performance on various clinical assessments, the presence of other biomarkers, and clinical outcomes were evaluated.

**Results:** Hyposmia was associated with reductions in striatal DaT binding and α-syn SAA positivity. MDS Prodromal PD Probability Scores, which incorporate DaT and olfactory function, predicted SAA positivity and phenoconversion. DaT positivity was much more common (80%) among phenoconverters (RBD-PC), than non-phenoconverters (10%). No significant motor or non-motor symptom differences were observed between the two groups at baseline, likely due to the small sample size.

**Conclusions:** α-syn SAA positivity, DaT positivity, and hyposmia are highly associated with each other. MDS Prodromal PD Probability scores may be useful predictors of near-term progression, and thus as stratification factors in clinical research study design.

---

REM Sleep Behavior Disorder (RBD) is a parasomnia characterized by dream enactment stemming from the loss of muscle atonia during REM sleep. RBD is one of the two main preclinical syndromes, along with hyposmia, of synucleinopathies, including Parkinson’s Disease (PD), Dementia with Lewy Bodies (DLB), and Multiple System Atrophy.^1^ Approximately 73.5% of individuals are diagnosed with one of the recognized clinical synucleinopathy syndromes within 12 years.^2^ Therefore, elucidating the factors contributing to the evolution of RBD to synucleinopathy syndromes with life-limiting motor, cognitive and/or autonomic manifestations is essential for developing preventative therapeutics aimed at mitigating the onset of PD. Biological markers, including striatal dopamine transporter (DaT) imaging, CSF synuclein aggregating activity (SAA), together with clinical characteristics, (hyposmia, clinical scales), are all potentially predictive measures of the progression of synucleinopathy in persons with RBD.

We recruited individuals with RBD, PD without RBD, PD with RBD (PD/RBD), and healthy controls (HC) to a study to investigate the potential role of and interactions between the gut microbiome and the immune system in the pathogenesis of the earliest stages of synucleinopathies. The results of the immunological aspects of this study are reported elsewhere (Zhang et al, submitted). The present report describes the clinical and biomarker characterization of the cohort and investigates the interplay among key PD clinical and biological markers including SAA positivity, olfactory function, and striatal dopamine receptor (DaT) imaging for nigrostriatal integrity. We hypothesized that RBD subjects manifesting hyposmia and/or reduced striatal DaT signal would exhibit a more severe clinical phenotype and have a higher likelihood of demonstrating synuclein aggregating activity in CSF, and a greater propensity to phenoconvert to PD or DLB in the near term.

## Methods

### Study Population

In all, we recruited persons with polysomnographically-proven RBD without REM sleep atonia (N = 36), Parkinson’s disease with (N = 15) and without concurrent RBD (n =18), as well as 15 healthy control subjects without evidence of neurological disease or other significant medical illnesses. Subjects in the Yale “RBD+” cohort were recruited using multiple sources, including Yale research registries, HIPAA-compliant medical records searches, outreach to local sleep clinics and Facebook posts. Inclusion/Exclusion criteria are shown in **Supplementary Table 1**. All subjects provided written or online informed consent to participate in this study. The study received necessary approvals from the Yale Institutional Review Board.

#### Clinical and Biomarker Assessments

Subjects completed a comprehensive battery of clinical assessments to assess motor, non-motor, and cognitive function assessments, generally spread across two or three virtual and in-person visits over the span of one month. All subjects underwent lumbar puncture with withdrawal of 30 mL of CSF. In addition, 150 mL of blood was drawn to support immunological and biomarker investigations. The clinical and biomarker assessments performed are summarized in **Table 1**.

**Table 1:** Clinical and Biomarker Assessments.

| **Non-Motor rating scales** |  |
| --- | --- |
| 1. | Olfactory testing (University of Pennsylvania Smell Identification Test) * |
| 2. | Montreal Cognitive Assessment (MoCA)* |
| 3. | Scale for Outcomes in Parkinson's Disease for Autonomic Symptoms (SCOPA-AUT) * |
| 4. | Movement Disorders Society-Unified Parkinson's Disease Rating Scale (MDS-UPDRS) Part I* |
| 5. | Movement Disorders Society-Prodromal Parkinson's Disease Likelihood Calculation * |
| 6. | Epworth Sleepiness Scale (ESS) |
| 7. | Non-Motor Symptoms Scale (NMSS) |
| 8. | REM Sleep Behavior Disorder Screening Questionnaire (RBDSQ) |
| 9. | Paris Arousal Disorders Severity Scale (PADSS) |
| 10. | Rome Constipation Criteria |
| 11. | Dietary Environmental Survey |
| **Motor rating scales** |  |
| 1. | Movement Disorders Society-Unified Parkinson's Disease Rating Scale (MDS-UPDRS) Parts II and III * |
| 2. | Brief Motor Questionnaire (BMQ) * |
| **Biomarker assessments** |  |
| 1. | Dopamine Transporter (DaT) Scan (RBD participants only) * |
| 2. | CSF Synuclein Aggregation Assay (SAA; Amprion) * |

*Assessments marked with an asterisk are presented and discussed in this paper.*

### Follow-up

RBD subjects were followed at one to two-year intervals to evaluate clinical status. The full results of these evaluations will be reported at a future date. Subjects who received a diagnosis of PD or DLB between study visits, or who were begun on antiparkinsonian medications or cholinesterase inhibitors were termed “phenoconverters.” It should be noted that this was a practical definition; we did NOT base our definition of phenoconversion on strict application of MDS-UPDRS criteria for PD^3^ or McKeith criteria for DLB^4^, since, as will be shown, there was considerable overlap in disease manifestations between the RBD and clinically manifest populations.

### Biomarker-Defined Axes

For the purposes of further exploration, we divided the RBD cohort along a set of 5 axes based on presence or absence of individual clinical or biological markers as described below. Along each axis subjects were compared across the remaining clinical and biomarker assessments and outcomes.

1. *<u>Olfactory Status (Hyposmic vs. Normosmic):</u>* RBD participants were retrospectively classified as hyposmic or normosmic groups based on their University of Pennsylvania Smell Identification Test (UPSIT) scores. Hyposmia was defined as a UPSIT score below the 15th percentile for age- and gender-matched norms.^5^
2. *<u>Dopamine Transporter SPECT:</u>* RBD participants were classified based on the age- and gender-specific percentile rank of the mean striatal specific binding ratio (SBR) as described under Methods above, resulting in 3 subgroups as follows: DaT-Positive (≤65th percentile; N = 7); DaT-Negative (≥ 80^th^ percentile; N= 17), and DaT-Intermediate (percentile >65 and < 80; N = 11).
3. *<u>SAA Status (SAA Positive vs. SAA Negative):</u>* RBD participants were retrospectively classified into SAA-positive and SAA-negative groups based on results of CSF SAA testing.

### Axes based on clinical outcomes

4. *<u>Prodromal PD Likelihood, based on the MDS Online Prodromal PD Calculator:</u>* RBD participants were categorized into high (≥ 80%, N = 26), intermediate (50–80%, N = 6), or low (<50%, N = 4) probability groups for having prodromal PD using the Movement Disorder Society’s online Prodromal PD calculator. Due to small sample sizes in the low and intermediate groups, these two were combined into a single low + intermediate category (N = 10).
5. *<u>Phenoconversion (Phenoconverters [PC] vs. Non-phenoconverters [nPC]):</u>* RBD participants who “phenoconverted” i.e., were diagnosed by the neurologist providing their care (generally not a member of the study team) with PD or DLB during the observation period following their baseline visit were classified as “phenoconverters” (PC).

### Statistical Analysis

The analysis herein focuses solely on the RBD cohort, examining the distribution of clinical and biomarker characteristics along each of the 3 biomarker axes with the goals of 1) defining dimensions that contribute to the utility of the MDS Prodromal PD Likelihood Ratio to predict the presence of synuclein pathology as reflected in the SAA or 2) the ability of the various biomarkers or the MDS Prodromal Score to predict clinically-defined phenoconversion in our cohort. Baseline descriptive statistics, including means and standard deviations, were calculated for demographic variables and all assessments. The distribution of the data for each variable of interest was first checked for normality using the Shapiro-Wilk test. Univariate associations between diagnostic subgroups and assessment scores were examined using chi-square tests for categorical variables. For continuous variables, parametric tests (t-tests and one-way ANOVAs) were used for normally distributed data, while non-parametric tests (Mann-Whitney and Kruskal-Wallis) were applied for non-normally distributed data. Statistical significance was determined with a two-tailed p-value threshold of 0.05. Analyses were performed using PRISM v10 or JMP v17. Given the exploratory nature of the analysis, no corrections have been made for multiplicity. Pairwise comparisons between groups were adjusted using the Steel-Dwass method if global tests yielded significant results.

### Data Availability

Data generated in this study were deposited to the Global Parkinson’s Genetics Program (GP2; https://gp2.org). GP2 is funded by the Aligning Science Across Parkinson’s (ASAP) initiative and implemented by The Michael J. Fox Foundation for Parkinson’s Research. Specifically, they are Tier 2 data from GP2 release 10.5281/zenodo.7904831. Tier 2 data access requires approval and a Data Use Agreement signed by your institution. Anonymized data, used and generated in this study are available alongside their persisent identifiers at [GP2 ID: YMS]. No code was generated for this study; all data cleaning, preprocessing, analysis, and visualization was performed using JMP Pro (v.17), OriginPro (v. 10) and PRISM (v.10).

A Key Resource Table can be found at the following DOI: 10.5281/zenodo.15685275.

## Results

We recruited a total of 84 subjects, 34 with RBD, 33 with PD, including 15 without and 18 with RBD, and 15 healthy controls (HC) between May 2021 and May 2024. Demographic and clinical characteristics of the 4 groups are detailed in **Table 2**. **Figure 1** demonstrates that motor function (MDS-UPDRS Part II and Part III scores) was most impaired in the PD/RBD group and least impaired in the RBD group, scores on non-motor assessments were similar across PD with and without RBD and RBD subjects. Interestingly a sub-group of the RBD cohort had the lowest MoCA scores among all the subjects. At this time, we cannot tell if subjects with lower MoCA scores in our cohort are at greater risk for cognitive, as opposed to motor decline.

**Figure 1:**
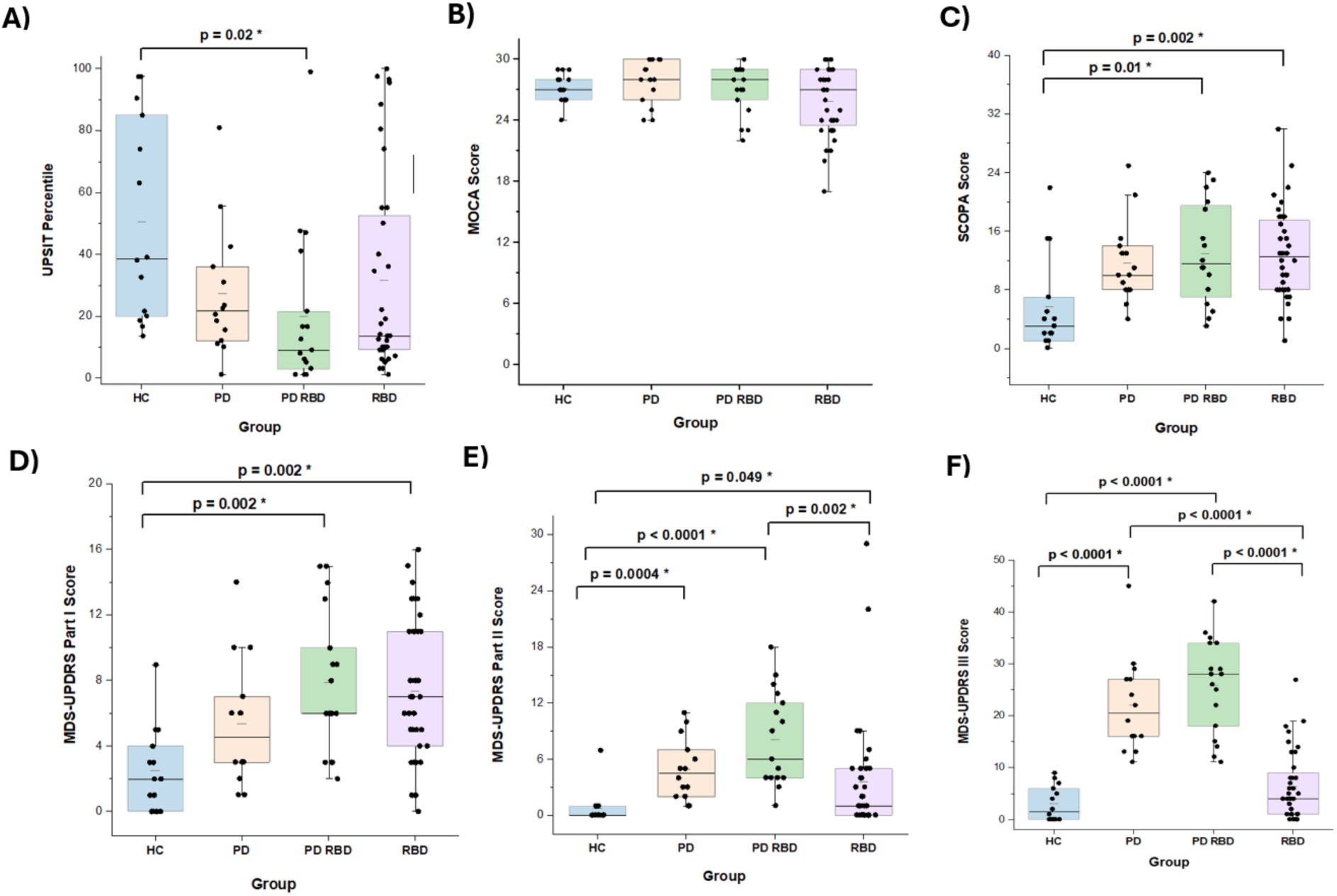
Clinical Features by Diagnostic Grouping. Box plots depict the median, interquartile range, and individual data points for each group. 1A. UPSIT percentiles. 1B. MoCA scores. 1C. SCOPA-AUT. 1D. MDS-UPDRS Part I. 1E. MDS-UPDRS Part II. 1F. MDS-UPDRS Part III.

**Table 2.** Association of Demographic and Clinical Variables with Hyposmia Status.

|  | RBD Hyposmic<br>N=20 | RBD Normosmic<br>N=16 | Hyposmic vs<br>Normosmic<br>p value |
| --- | --- | --- | --- |
| Age | 71.1 ± 4.3 | 66.1 ± 6.9 | 0.11 |
| Male (n, %) | 16 (80.0%) | 9 (56.2%) | 0.77 |
| White (n, %) | 20 (100%) | 16 (100%) | 0.89 |
| **Non-Motor Assessments** |  |  |  |
| MOCA | 25.9 ± 2.9 | 25.9 ± 3.9 | 0.50 |
| UPSIT Percentile | 8.7 ± 3.7 | 60.1 ± 30.3 | **<0.0001** |
| SCOPA-AUT | 12.7 ± 4.5 | 13.1 ± 8.1 | 0.29 |
| UPDRS Part I Scores | 7.2 ± 4.3 | 7.5 ± 4.5 | >0.99 |
| **Motor Assessments** |  |  |  |
| BMQ | 0.9 ± 1.8 | 1.8 ± 2.1 | 0.80 |
| UPDRS Part II Scores | 2.7 ± 6.4 | 4.7 ± 5.5 | 0.22 |
| UPDRS Part III Scores | 7.0 ± 7.7 | 5.6 ± 5.1 | >0.99 |
| **DaT Data** |  |  |  |
| Mean Striatal SBR Percentile | 70.0 ± 22.8 | 88.9 ± 25.8 | **0.03** |
| Mean Striatal SBR | 1.7 ± 0.5 | 2.2 ± 0.6 | **0.01** |
| DaT Positivity<br>(Mean striatal SBR percentile of ≤ 65%) | 6 (30%) | 1 (6.3%) | 0.06 |
| **SAA Data** |  |  |  |
| SAA Positivity | 20 (100%) | 6 (37.5%) | **<0.0001** |
| **MDS-Prodromal PD Calculator** |  |  |  |
| Probability of Prodromal PD | 0.9 ± 0.08 | 0.7 ± 0.3 | **0.005** |
| Prodromal PD Positivity (Estimated probability ≥ 80%) | 18 (90%) | 8 (50%) | **0.009** |

Data are presented as sample size (percentage) for categorical variables and mean ± standard deviation for continuous variables. Bolded p-values indicate significance.

### Axis 1: Olfactory Status

Sixteen RBD subjects were classified as normosmic and 20 as hyposmic (**Table 2)**. The normosmic group tended to be younger, though not statistically significantly so. There were no differences in any of the motor, non-motor or cognitive outcome assessments between hyposmic and normosmic subjects other than, as might be expected, UPSIT scores.

Overall, the hyposmic subjects had lower DaT binding. 20 of the hyposmic subjects were SAA-positive, as opposed to only 37.5% of the normosmic group. Finally, as expected since DaT positivity and hyposmia weigh heavily in the calculation of the MDS Prodromal PD Calculator Likelihood Ratio (LR) score, a higher percentage of hyposmic than normosmic subjects were classified by the online calculator has having an 80% or greater probability of having prodromal PD.

### Axis 2: DaT Binding Ratios

As shown in **Table 3**, 7 subjects were classified as DaT-positive, 17 as DaT-negative, and 11 as DaT-intermediate. There were no differences in mean age between the 3 groups. DaT-positive and DaT-intermediate groups had lower mean UPSIT percentile scores than did DaT-negative subjects. There were no differences in motor, other non-motor or clinical outcome assessment scores between the groups. SAA positivity was higher in the DaT-positive and DaT-intermediate groups compared with the DaT-negative group, but this trend was not statistically significant. As expected, mean MDS Prodromal Probability scores were highest among the DaT-positive group, and there was a non-significant trend in the proportion of subjects achieving prodromal probability scores ≥ 80% for the DaT-positive, DaT-intermediate, and DaT-negative groups, respectively. This result is to be expected, since DaT positivity is a major driver of the Prodromal Likelihood Ratio (https://www.movementdisorders.org/Prodromal-PD-Calculator.htm).

**Table 3:**
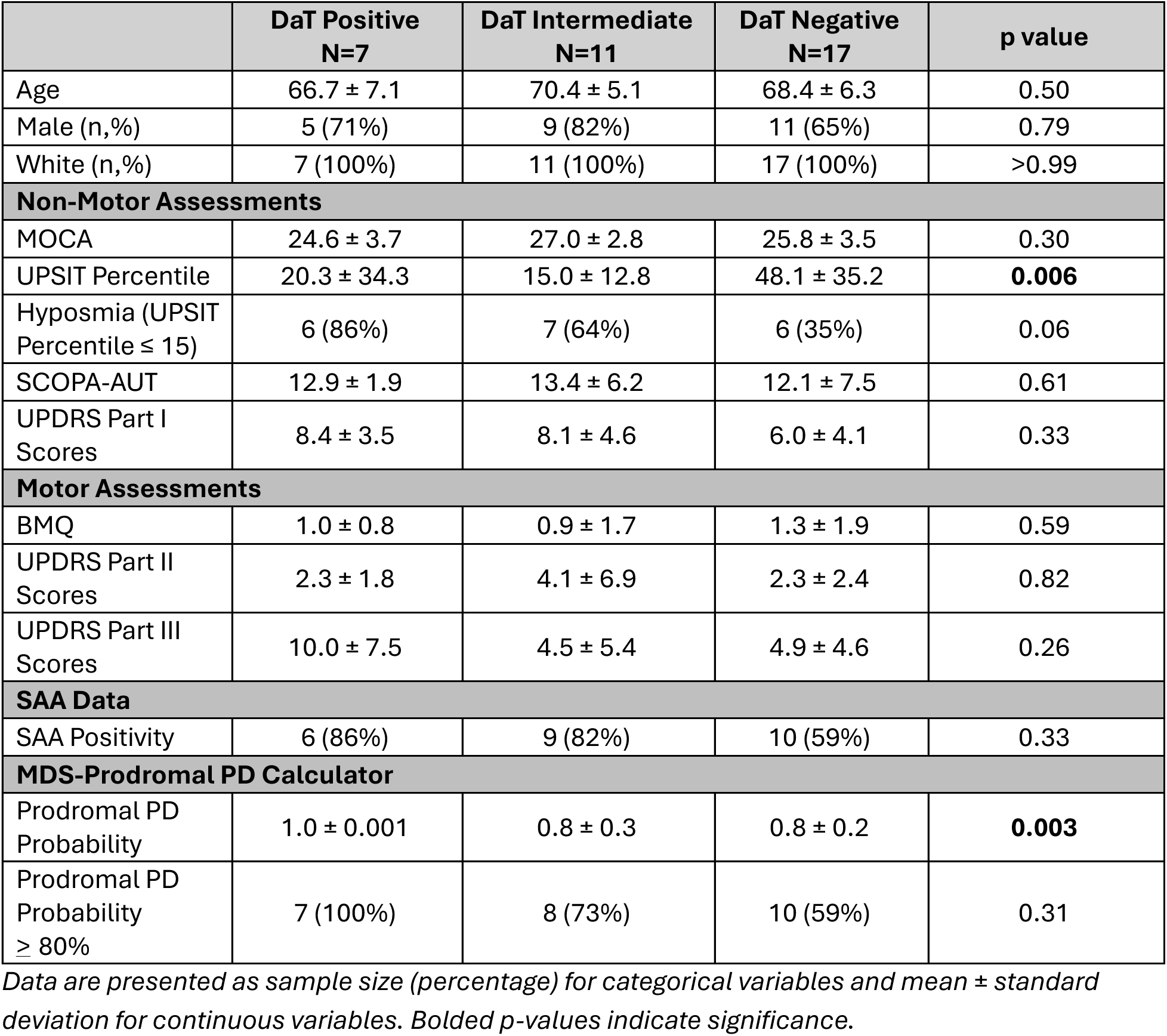
Association of Demographic and Clinical Variables with DaT Binding.

|  | **DaT Positive**<br>**N=7** | **DaT Intermediate**<br>**N=11** | **DaT Negative**<br>**N=17** | **p value** |
| --- | --- | --- | --- | --- |
| Age | 66.7 ± 7.1 | 70.4 ± 5.1 | 68.4 ± 6.3 | 0.50 |
| Male (n,%) | 5 (71%) | 9 (82%) | 11 (65%) | 0.79 |
| White (n,%) | 7 (100%) | 11 (100%) | 17 (100%) | >0.99 |
| **Non-Motor Assessments** |  |  |  |  |
| MOCA | 24.6 ± 3.7 | 27.0 ± 2.8 | 25.8 ± 3.5 | 0.30 |
| UPSIT Percentile | 20.3 ± 34.3 | 15.0 ± 12.8 | 48.1 ± 35.2 | **0.006** |
| Hyposmia (UPSIT Percentile ≤ 15) | 6 (86%) | 7 (64%) | 6 (35%) | 0.06 |
| SCOPA-AUT | 12.9 ± 1.9 | 13.4 ± 6.2 | 12.1 ± 7.5 | 0.61 |
| UPDRS Part I Scores | 8.4 ± 3.5 | 8.1 ± 4.6 | 6.0 ± 4.1 | 0.33 |
| **Motor Assessments** |  |  |  |  |
| BMQ | 1.0 ± 0.8 | 0.9 ± 1.7 | 1.3 ± 1.9 | 0.59 |
| UPDRS Part II Scores | 2.3 ± 1.8 | 4.1 ± 6.9 | 2.3 ± 2.4 | 0.82 |
| UPDRS Part III Scores | 10.0 ± 7.5 | 4.5 ± 5.4 | 4.9 ± 4.6 | 0.26 |
| **SAA Data** |  |  |  |  |
| SAA Positivity | 6 (86%) | 9 (82%) | 10 (59%) | 0.33 |
| **MDS-Prodromal PD Calculator** |  |  |  |  |
| Prodromal PD Probability | 1.0 ± 0.001 | 0.8 ± 0.3 | 0.8 ± 0.2 | **0.003** |
| Prodromal PD Probability ≥ 80% | 7 (100%) | 8 (73%) | 10 (59%) | 0.31 |

Data are presented as sample size (percentage) for categorical variables and mean ± standard deviation for continuous variables. Bolded p-values indicate significance.

### Axis 3: SAA-Positive vs SAA-Negative

26 of the RBD subjects demonstrated SAA positivity in their CSF; 10 did not (**Table 4**). SAA-positive individuals had a lower mean UPSIT percentile score; 20 (77%) met criteria for hyposmia. Mean striatal SBR scores were modestly but not significantly lower in the SAA-positive group, driven by the fact that six of the SAA-positive subjects had SBRs less than the 65^th^ percentile, compared with only one SAA-negative subject. The SAA-positive group had a greater mean MDS-UPDRS prodromal calculator probability score than the SAA-negative group, with 77% of SAA-positive and just 50% of SAA-negative subjects achieving a prodromal PD probability of 80% or greater.

**Table 4.** Association of Demographic and Clinical Variables with SAA Status.

|  | **SAA**<br>**Positive**<br>**N= 26** | **SAA**<br>**Negative**<br>**N=10** | **SAA Positive vs.**<br>**Negative**<br>**p value** |
| --- | --- | --- | --- |
| Age | 70.4 ± 4.3 | 64.9 ± 8.3 | **0.01** |
| Male (n,%) | 19 (73%) | 6 (60%) | 0.45 |
| White (n,%) | 26 (100%) | 10 (100%) | 0.99 |
| **Non-Motor Assessments** |  |  |  |
| MOCA | 26.5 ± 3.0 | 24.2 ± 3.6 | **0.06** |
| UPSIT Percentile | 18.2 ± 23.1 | 66.0 ± 29.1 | **<0.0001** |
| Hyposmia (UPSIT<br>Percentile ≤ 15) | 20 (77%) | 0 (0%) | **<0.0001** |
| SCOPA-AUT | 12.9 ± 5.6 | 12.7 ± 8.1 | 0.88 |
| UPDRS Part I Scores | 7.0 ± 4.2 | 8.5 ± 5.0 | 0.35 |
| **Motor Assessments** |  |  |  |
| BMQ | 1.0 ± 1.8 | 2.3 ± 2.2 | **0.08** |
| UPDRS Part II Scores | 2.8 ± 5.8 | 5.8 ± 6.3 | **0.02** |
| UPDRS<br>Part III Scores | 6.3 ± 7.0 | 6.7 ± 6.1 | 0.72 |
| **DaT Data** |  |  |  |
| Mean Striatal<br>SBR Percentile | 75.6 ± 24.4 | 86.2 ± 28.4 | 0.28 |
| Mean Striatal SBR | 1.8 ± 0.6 | 2.2 ± 0.7 | 0.16 |
| DaT Positivity (mean<br>striatal SBR<br>percentile of ≤ 65%) | 6 (23%) | 1 (10%) | 0.27 |
| **MDS-Prodromal PD Calculator** |  |  |  |
| Prodromal PD<br>Probability | 0.9 ± 0.2 | 0.7 ± 0.3 | **0.045** |
| Prodromal PD<br>probability ≥ 80% | 20 (77%) | 5 (50%) | **0.08** |

Data are presented as sample size (percentage) for categorical variables and mean ± standard deviation for continuous variables. Bolded p-values indicate significance.

### Axis 4: Prodromal Probability Likelihood Ratio Scores

It should be kept in mind that the algorithm that calculates the MDS Prodromal Probability score heavily weights age, male gender, DaT positivity, and hyposmia. We nonetheless calculated Prodromal Probability Likelihood Ratios for subjects in our RBD cohort and grouped them into Low (<50%; N = 4), Intermediate (50-80%; N = 6) and High (≥ 80%; N = 26) groups. Because of the limited number of subjects in each of the Low and Intermediate probability groups, these were combined into a single group for analysis. **Table 5** displays the data for the Low and Intermediate, as well as the pooled Low + Intermediate groups, but comparisons are limited to the High vs Low + Intermediate groups only. As expected, high prodromal PD probability scores were related to lower mean UPSIT percentile scores and mean striatal SBR percentiles as well as higher SCOPA-AUT scores and MDS-UPDRS Part 1 and 3 scores. However, cognition, as reflected by total MoCA scores, was similar between the groups as reflected by MoCA scores. Of note, 81% of the High probability group was SAA-positive, compared to only 50% of the Low + Intermediate group. This observation raises the intriguing question of whether SAA-positivity occurs before dopaminergic neuron degeneration becomes fully evident as assessed by current DAT-SPECT methods.

**Table 5.** Association of Demographic and Clinical Variables with Prodromal PD Likelihood Ratio Probabilities.

|  | Low<br>(<50%)<br>N=4 | Intermediate<br>(50-80%)<br>N=6 | Low +<br>Intermediate<br>(0-80%)<br>N=10 | High<br>(>80%)<br>N=26 | Low +<br>Intermediate<br>vs. High<br>p value |
| --- | --- | --- | --- | --- | --- |
| Age | 66.7 ± 7.8 | 68.0 ± 8.2 | 67.5 ± 7.6 | 69.4 ± 5.5 | 0.41 |
| Male (n,%) | 3 (75%) | 2 (33%) | 5 (50%) | 20 (77%) | 0.22 |
| **Race** |  |  |  |  |  |
| White (n,%) | 4 (100%) | 6 (100%) | 10 (100%) | 26 (100%) | 0.99 |
| **Non-Motor Assessments** |  |  |  |  |  |
| MOCA | 25.7 ± 4.4 | 26.3 ± 2.6 | 26.1 ± 3.2 | 25.8 ± 3.4 | 0.88 |
| UPSIT<br>Percentile | 65.2 ± 31.7 | 54.6 ± 37.0 | 58.8 ± 33.6 | 21.0 ± 26.1 | **0.002** |
| Hyposmia<br>(UPSIT<br>Percentile ≤ 15) | 0 (0%) | 2 (33%) | 2 (20%) | 18 (69%) | **0.008** |
| SCOPA-AUT | 10.7 ± 7.9 | 8.8 ± 5.5 | 9.6 ± 6.2 | 14.1 ± 5.9 | **0.05** |
| UPDRS Part I | 7.0 ± 5.9 | 4.3 ± 4.7 | 5.4 ± 5.1 | 8.1 ± 3.9 | 0.06 |
| **Motor Assessments** |  |  |  |  |  |
| BMQ | 1.2 ± 1.5 | 1.2 ± 2.0 | 1.2 ± 1.7 | 1.4 ± 2.1 | 0.89 |
| UPDRS Part II | 7.7 ± 10.4 | 1.3 ± 2.0 | 3.9 ± 7.0 | 3.5 ± 5.8 | 0.51 |
| UPDRS Part III | 1.5 ± 1.7 | 1.5 ± 1.6 | 1.5 ± 1.7 | 8.3 ± 6.9 | **0.001** |
| **DaT** |  |  |  |  |  |
| Mean Striatal<br>SBR Percentile | 83.2 ± 22.6 | 96.0 ± 24.8 | 90.9 ± 23.6 | 71.1 ± 28.3 | 0.06 |
| Mean Striatal<br>SBR | 2.0 ± 0.6 | 2.4 ± 0.7 | 2.2 ± 0.6 | 1.8 ± 0.6 | **0.05** |
| DaT Positivity<br>(Mean striatal<br>SBR percentile<br>of ≤ 65%) | 0 (0%) | 0 (0%) | 0 (0%) | 7 (27%) | 0.14 |
| **SAA** |  |  |  |  |  |
| SAA Positivity | 1 (25%) | 4 (67%) | 5 (50%) | 21 (81%) | 0.10 |

Data are presented as sample size (percentage) for categorical variables and mean ± standard deviation for continuous variables. Bolded p-values indicate significance.

Because of the individual associations between SAA positivity and DaT and olfactory function, we assessed overlap between DaT, SAA, and olfactory testing in our 35 RBD participants (**Figure 2)**. All of the hyposmic individuals were also SAA positive, reinforcing the strong correlation between SAA positivity and olfactory deficits and underscoring the potential for combining RBD and hyposmia as screening criteria to identify a population with high probability of having an underlying synucleinopathy for future secondary prevention clinical trials.

**Figure 2.**
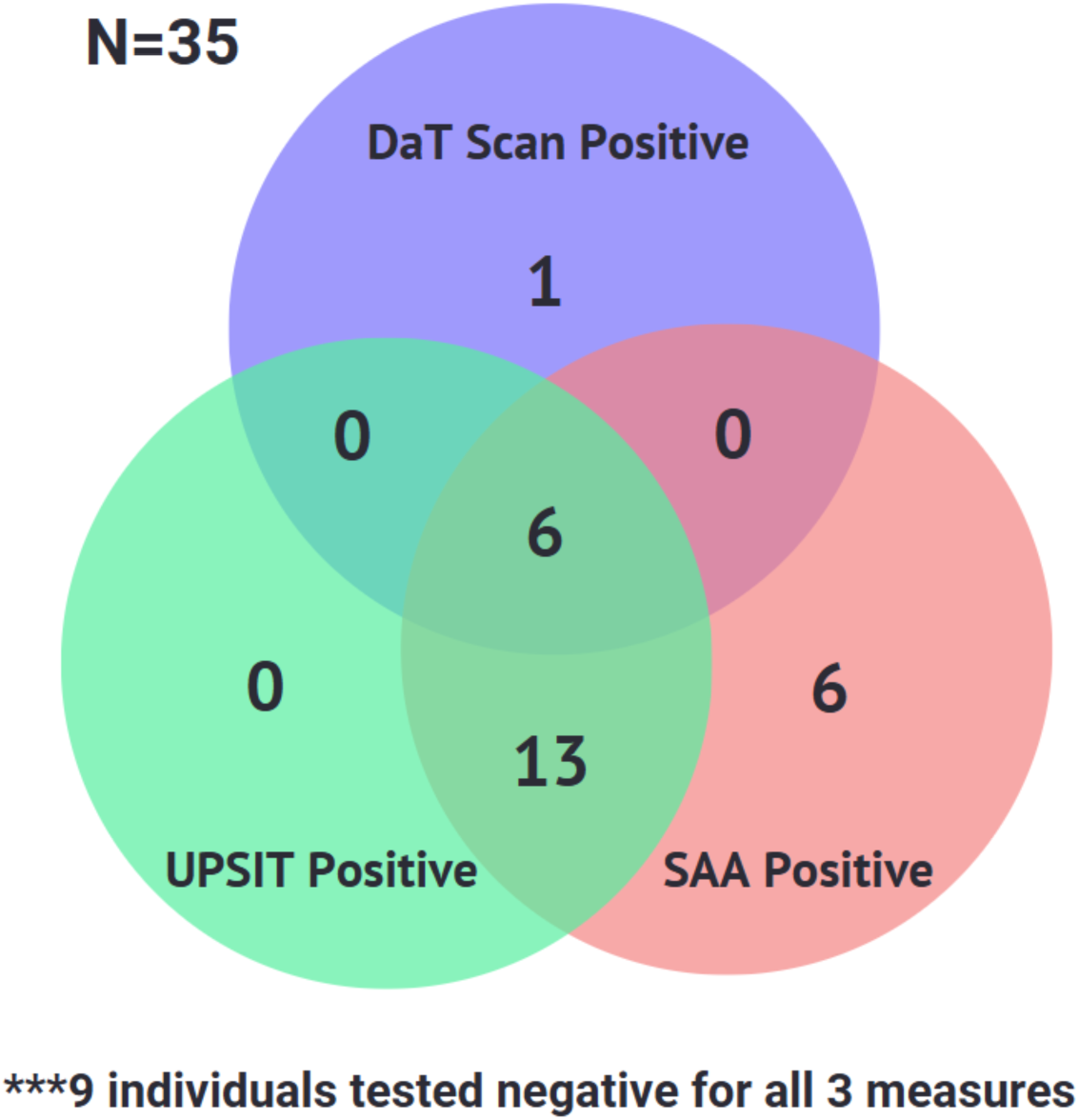
Overlap of DaT, SAA, and Olfactory Testing (UPSIT) in RBD subjects. The numbers within each region represent the count of individuals meeting the respective criteria.

### Axis 5: Phenoconverters and Non-Phenocoverters

During the 4-year period during which the cohort was being assembled, 6 RBD subjects were diagnosed by outside clinicians with either PD (5 subjects) or DLB (1 subject) (**Table 6**). These diagnoses were rendered by outside physicians and not by the study team; no formal diagnostic criteria were used to establish or confirm the diagnoses. Here we review the data from the study baseline visit only for these individuals. RBD-PC and RBD-nPC subjects did not difer significantly in age, gender distribution, non-motor, cognitive or motor assessment scores at baseline. However, mean scores on the MoCA were numerically lower, and scores on the BMSQ and all 3 parts of the MDS-UPDRS were numerically higher, in the PC group at the baseline visit. The lack of nominal statistical significance for these measures is likely a reflection of the small sample size in this study. The PC group had a lower mean striatal SBR percentile and higher DaT positivity rates than the nPC group. However, the proportion of SAA-positive subjects was similar between the two groups, though slightly higher in the PC group, again consistent with the concept that SAA positivity precedes substantia dopamine neuron loss. 100% of the phenoconverters had an MDS-UPDRS prodromal probability score ≥ 80% compared with 67% in the nPC group, with a greater mean LR probability in the PC group.

**Table 6.** Demographic, Clinical and Biomarker Characteristics of Phenoconverters (PC and non-Phenoconverters.

|  | **RBD-nPC**<br>**N=30** | **RBD-PC**<br>**N= 6** | **nPC vs PC**<br>**p value** |
| --- | --- | --- | --- |
| Age | 68.9 ± 1.0 | 68.5 ± 3.7 | 0.99 |
| Male | 21 (70.0%) | 4 (66.7 %) | 0.99 |
| White | 30 (100%) | 6 (100%) | 0.99 |
| **Non-Motor Assessments** |  |  |  |
| MOCA | 26.4 ± 3.2 | 23.3 ± 2.9 | 0.23 |
| UPSIT Percentile | 33.3 ± 32.2 | 22.7 ± 36.8 | 0.71 |
| Hyposmia (UPSIT Percentile ≤ 15) | 15 (50%) | 5 (83%) | 0.20 |
| SCOPA-AUT | 12.4 ± 6.5 | 15.3 ± 4.5 | 0.64 |
| UPDRS Part I Scores | 6.8 ± 4.0 | 10.3 ± 5.2 | 0.50 |
| **Motor Assessments** |  |  |  |
| UPDRS Part II Scores | 3.0 ± 4.5 | 6.5 ± 11.2 | 0.97 |
| UPDRS Part III Scores | 5.3 ± 5.2 | 11.8 ± 10.4 | 0.63 |
| BMQ | 1.2 ± 1.7 | 2.0 ± 3.1 | 0.99 |
| **DaT Data** |  |  |  |
| Mean Striatal SBR Percentile | 84.5 ± 21.2 | 43.8 ± 23.8 | **0.0004** |
| Mean Striatal SBR | 2.1 ± 0.5 | 1.7 ± 0.5 | **0.002** |
| DaT Positivity (mean striatal SBR percentile ≤ 65%) | 3 (10%) | 5 (80%) | **0.003** |
| **SAA Data** |  |  |  |
| SAA Positivity | 19 (70%) | 5 (83%) | >0.99 |
| **MDS-Prodromal PD Calculator** |  |  |  |
| Prodromal PD Probability | 0.8 ± 0.3 | 1.0 ± 0.001 | **0.0004** |
| Prodromal PD Probability ≥ 80% | 20 (67%) | 6 (100%) | 0.48 |

Data are presented as sample size (percentage) for categorical variables and mean ± standard deviation for continuous variables. Bolded p-values indicate significance.

## Discussion

We evaluated non-motor and motor symptoms as well as DaT, SAA, and UPSIT biomarkers across healthy controls, RBD subjects, PD/RBD subjects, and PD subjects. We characterized our RBD population along 5 axes: hyposmic versus normosmic individuals; SAA-positive versus SAA-negative individuals; DaT positive, intermediate, and negative individuals; and individuals with high versus low/intermediate prodromal PD likelihood ratios.

The demographics and clinical characteristics of the cohort we recruited are typical of others reported in the literature, including PPMI and ICEBERG ^6^, and the North American Prodromal Synucleinopathy (NAPS)^7^ multicenter cohorts (**Supplementary Table 1)**. 56% of our RBD cohort was hyposmic, half our RBD subjects had reduced DaT binding in the striatum, and 72% of our RBD subjects’ CSF tested positive in the SAA assay. There was considerable overlap among these markers. All hyposmic subjects were SAA-positive, 77% of SAA-positive subjects were also hyposmic, and 6 of 7 subjects with DaT SBR percentiles in the PD range were both hyposmic and SAA-positive.

When comparing the subgroup analyses against previously conducted studies, similarities can be identified. Evaluation of olfactory data across studies indicates both similarities and differences. While Lee et al. found that olfactory impairment in RBD was correlated with reduced tracer uptake in the caudate as well as the anterior and posterior putamen^8^, the present study identified similar findings in which hyposmic RBD subjects had reduced mean striatal SBR percentiles and mean striatal SBR values when compared to normosmic subjects (Table 2). However, Lee et al. found increased constipation and deficits in urinary and sexual function in hyposmic subjects, but not those without; the present study found no significant changes in any of the aforementioned symptoms between hyposmic versus normosmic RBD groups (Table 2). Like the present study, they found insignificant cognitive differences between hyposmic versus normosmic groups. Alternatively, Kang et al. found that RBD hyposmics exhibited lower cognition than RBD normosmics.^9^

Similarly to the findings of Siderowf et al.^10^, hyposmic subjects in our cohort had significantly lower mean striatal SBR percentiles and mean striatal SBR values than did normosmic subjects (Table 2). Additionally, more hyposmic subjects were SAA-positive than were normosmic subjects, with all hyposmic subjects showing SAA positivity (Table 2). Conversely, no SAA-negative subjects were found to be hyposmic. These observations are consistent with a model in which pathological changes appear to precede clinical symptoms, as SAA-positive and DaT-positive individuals show no significant clinical differences from their negative counterparts aside from lower UPSIT percentiles. As there were no UPSIT positive subjects who did not also show either SAA positivity or SAA and DaT positivity, we hypothesize that SAA positivity likely precedes both hyposmia and DaT positivity, with hyposmia emerging next and DaT positivity emerging last.

These observations are consistent with a model in which pathological changes appear to precede clinical symptoms, as SAA-positive and DaT-positive individuals show no significant clinical differences from their negative counterparts aside from lower UPSIT percentiles. As there were no UPSIT positive subjects who did not also show either SAA positivity or SAA and DaT positivity, we hypothesize that SAA positivity likely precedes both hyposmia and DaT positivity, with hyposmia emerging next and DaT positivity emerging last.

DaT positivity and hyposmia all correlated to higher probabilities of prodromal PD according to the prodromal PD calculator^11^. This is to be expected, as both of these biomarkers are weighted heavily in the likelihood ratio calculation. Not surprisingly, the proportion of SAA-positive individuals increased in step with estimated prodromal PD probability in our RBD population. Our RBD cohort showed a strong overlap between UPSIT and SAA positivity as well as some overlap between DaT, SAA, and UPSIT positivity (**Figure 2**).

Within our small RBD cohort, six subjects clinically phenoconverted to PD or DLB during a relatively short period of observation. Various external studies reflected similar findings to our study regarding PCs. ^2,12,13,14^ In all cohorts, a greater percentage of PCs had abnormal DaT scans than did nPCs. In our RBD cohort along with Postuma et al. and Arnaldi et al., a greater percentage of PCs were hyposmic than were nPCs. ^2,12^ Iranzo et al. showed a higher UPSIT percentile average and lower percentage of SAA-positive subjects in their nPC group, reflecting our results.^13^ All PCs were identified by the MDS Prodromal PD Calulator as having prodromal PD with >80% probability, while only 67% of nPC subjects were identified as having prodromal PD. Similar results were found by Postuma et al. and Iranzo et al. in which a greater percentage of PCs were identified as having prodromal PD than were nPCs.^2,13^ PCs in our cohort tended to have lower MoCA and higher SCOPA-AUT and MDS-UPDRS Parts II and III scores with more impairment in non-motor function. Postuma et al. paralleled our cohort findings for MDS-UPDRS II and III scores ^2^ while Arnaldi et al. paralleled our cohort findings for MDS-UPDRS III and MoCA scores.^12^

Interestingly, baseline motor (MDS-UPDRS Parts II and III) scores did not difer between eventual phenoconverters and those who did not develop motor or cognitive impairment during the period of observation. The pattern we observed is consistent with the hypothesis that within the RBD population, hyposmia and autonomic dysfunction appear to mark individuals for more rapid progression towards motor and/or cognitive-manifest synucleinopathy disorders. SAA positivity appears to precede DaT positivity, and finally cognitive and autonomic dysfunction, together with subtle motor dysfunction, immediately precede the onset of the recognized motor and/or cognitive syndromes.

On the other hand, in our cohort, non-motor function, as assessed by MDS-UPDRS Part 1 and SCOPA-AUT scores, was similarly impaired in our RBD subjects as in those diagnosed with PD who also experienced RBD. Neither the presence of hyposmia, a DaT deficit nor SAA positivity appeared to influence the degree of non-motor dysfunction. However, SCOPA-AUT scores were significantly higher among subjects who “phenoconverted,” and there was also a trend towards higher MDS-UPDRS Part 1 scores in this group (**Table 6**).

One question raised by our data is whether the pathogenic mechanisms underlying progression to PD from hyposmia and RBD are independent, additive or synergistic. In the PARS study, significantly more hyposmic RBD subjects showed DaT positivity than normosmic subjects^15^, and hyposmic individuals were subsequently diagnosed with PD in higher proportion than non-hyposmics.^16^ Dusek et al. showed that RBD subjects with lower DaT binding in the putamen had higher UPDRS part III scores, worse non-motor function, and higher prodromal PD likelihood ratios than counterparts with intermediate or normal DaT binding.^17^

Our study has several strengths. It features a comprehensive dataset that assesses non-motor/motor symptom and biomarker measures. All RBD cases were PSG-confirmed as well. We assessed both motor and non-motor symptoms using a battery of validated instruments. Importantly, we compared estimated probability of individuals truly having prodromal synucleinopathy to their SAA and other marker status.

Limitations of this study include the small sample size (n = 84), limited ancestral variation, with a predominantly white study cohort, and a lack of DaT imaging for PD and HC participants. Sex distribution was imbalanced as the cohort contained more males than females; however, this can be attributed to the fact that male sex is one of the strongest risk factors for RBD.^18^ Additionally, motor symptoms were assessed using the MDS-UPDRS, which can have sensitivity in early stages of disease.^19^ Activity monitoring devices can be used in future studies to provide more accurate and longitudinal motor symptom data.

Lastly, phenoconversion was determined by accepting diagnoses from providers outside the study. Though this allowed for a practical definition of phenoconversion, we must acknowledge our lack of prospective application of formal PD^3^ and DLB^4^ diagnostic criteria to validate the occurrence of phenoconversion. However, given the recent emphasis on and evolution of criteria for the staging continuity of synucleinopathy syndromes, we elected to refrain from the use of formal definitions.

In summary, our data are consistent with the construct that that appearance of pathological α-synuclein as measured by SAA precedes loss of dopaminergic nerve terminals, which is in turn followed by precede clinical signs and symptoms in RBD and PD^10^, though these changes vary among patient populations and do not necessarily appear in a linear progressive fashion, as shown in previous studies.^6^ SAA positivity, DaT positivity, and hyposmia are highly associated with each other. When encountered in individuals with RBD, hyposmia is a potential marker of nearer-term progression and phenoconversion.^20^ Finally, the MDS Prodromal Likelihood Score, which takes into account, among other things, age, gender, exposures, clinical status, DaT binding and hyposmia but not SAA, is a potentially useful predictor of SAA positivity and may be of use as a screening tool in future clinical trials.

## Data Availability

All data produced in the present study are available upon reasonable request to the authors

## Acknowledgments

We would like to thank all our RBD participants and their care partners who took time to participate in our study. We would also like to thank Drs. Kenny Susenko, Katy Zuchovsky, Sarah Mancone, and Olivia Gruder, who performed patient evaluations and lumbar punctures over the course of the study.

## Author Contributions

(1) Research Project: A. Conception, B. Organization, C. Execution; (2) Statistical Analysis: A. Design, B. Execution, C. Review and Critique; (3) Manuscript Preparation: A. Writing of the First Draft, B. Review and Critique.

V.L.R.: 1A, 1B, 1C, 2A, 2B, 2C, 3A, 3B

S.E.: 1C, 2C, 3A, 3B

E.R.: 1B, 1C, 2C, 3B

A.B.: 1C,3B

K.M.: 1B, 1C,3B

C.C: 1B, 1C,3B

S.P.: 1C,3B

J.S.: 1C, 2C, 3B

Y.H.: 2A, 2B, 2C, 3B

B. B.K.: 1C,3B

J.C.: 1A, 1B, 1C, 2A, 2B, 2C, 3A, 3B

## Financial Disclosure for the previous 12 months

Jesse Cedarbaum is supported by a grant from The Marcus Foundation (Grant # AWD0011411) and has received consulting fees from VanquaBio, Inc. and Merck KGA. Brian Koo is supported by The Program Restless Legs Syndrome Foundation Research Grant, “Cerebrospinal Fluid Melanocortins and Endorphin in Restless Legs Syndrome Related Augmentation”, and by the Stanley Center # 6910148-5500001534 (Broad Institute) Schizophrenia Spectrum Biomarkers Consortium. Yuan Huang is supported by NIH grant 5R01AG063946 and the CHDI Foundation: Modeling Early Natural History of HD. John Seibyl receives funding from the Michael J Fox Foundation (Grant # MJF-025524)

