## Supplement for "Characterizing Parkinson’s Disease Clinical and Biomarker Interactions in REM Sleep Behavior Disorder"

Supplementary Table 1.  Inclusion/Exclusion Criteria 
Inclusion Criteria:
Exclusion Criteria: 
1. Adult male or female
2. Age ≥ 40
3. English Speaking
4. Able and willing to provide informed consent
5. Repeated episodes of sleep-related vocalization and/or complex motor behaviors. 
6. Polysomnographic recording demonstrates REM sleep without atonia. 
1. An identified secondary cause of RBD, such as antidepressant medications or a diagnosis of narcolepsy. 
2. Personal history of any neurological disorder: Neurological examination must be performed to exclude parkinsonism (according to MDS PD criteria) and clinically evident dementia.
3. Known immune deficiency disorder
4. Solid or hematological malignancy other than in-situ carcinoma of the cervix or basal cell carcinoma of the skin
5. Known blood clotting disorder
6. Short bowel syndrome, ileostomy, pr colostomy
7. History of vagotomy
8. Pregnancy or actively breast feeding
9. Taking certain drugs that interfere with the interpretation of DaT scan
10. Receiving chemotherapy agents and/or immunosuppressant agents
11. Taking anticoagulant or antiplatelet medication. Low-dose (100 mg/day) aspirin is allowed.
12. Treatment of thyroid disease with radioactive iodine.


Supplementary Table 2: Comparison of Yale RBD+ Cohort to PPMI, ICEBERG and Oxford Discovery Populations.

Table modified after Di Folco et al6 Abbreviations: SBR = striatal binding ratio, y = years. Sources: PPMI, ICEBERG: DiFolco et al 2023; Oxford: Barber et al 2017. * Due to lack of motor symptom laterality in most RBD participants, SBR of Yale RBD+ Cohort is reported as mean of right and left for each basal ganglia region. **contra = contralateral (to side of worst symptoms)


Characterizing PD Clinical and Biomarker Interactions in RBD										Reddy et al
